# Long-range local influenza forecasts via distributed syndromic monitoring: preliminary results

**DOI:** 10.1101/2020.06.07.20078956

**Authors:** B.D. Dalziel, S.D. Chamberlain, C.A. Ariza, A.L. Daitch, P.P. Philips, I. Singh

## Abstract

Forecasting influenza primes public health systems to respond, reducing transmission, morbidity and mortality. Most influenza forecasts to date have, by necessity, relied on spatially course-grained data (e.g. state-or country-level incidence), and have operated at time horizons of 12 weeks or less. If influenza outbreaks could be predicted farther in advance and with increased spatial precision, then limited public health resources could be adaptively managed to minimize spread and improve health outcomes. Here, we use real-time syndromic data from a distributed network of thermometers to construct city-specific forecasts of influenza-like illness (ILI) with a horizon of 30 weeks. Daily geolocated ILI data from the network allows for estimates of recurrent city-specific patterns in ILI transmission rates. These “transmission templates” are used to parameterize an ensemble of ILI forecasts that differ randomly in three parameters, representing city- and season-specific rates of susceptible depletion and reporting, as well as differences in influenza season onset timing. For nine cities across the US, the best-in-hindsight model matches the observed data, and the best forecast variants can be identified in the early season.

## Introduction

Timely, location-specific forecasts of influenza-like illness (ILI) can enable more effective public health responses. These have the potential to reduce the size and impact of epidemics, for instance by prepositioning personnel, equipment and supplies to boost surge capacity, or by targeting vaccination or public health messaging campaigns to predicted transmission hotspots. The epidemic models that power these forecasts also allow inference into transmission dynamics, further enhancing predictive understanding of how drivers of transmission, including climate and demography, interact (*1*). These findings can inform public policy, and aid in monitoring and predicting the global circulation and evolution of influenza, with applications including vaccine development (*2*). Forecasting influenza also offers a unique window into the predictability of complex adaptive systems across scales, from viral interactions with host immune responses, to global patterns in the emergence and spread of novel strains (*2, 3*).

Influenza transmission is impacted by climatic conditions, especially specific humidity (*4*), and may be affected by population size, density and socioeconomic conditions (*1*). As these drivers vary geographically and seasonally, they combine to produce different patterns of transmission across space and time. Influenza transmission occurs on a spatial scale of meters (e.g. a significant fraction of transmission probably occurs at ranges of < 1m) (*5*) and a temporal scale of hours (e.g. a significant fraction of transmission probably occurs with 24-48 hours) (*6*). Leveraging incidence data closer to the scale of transmission may improve forecasts, providing more accurate and precise measurements of transmission dynamics.

Here we leverage city-specific data on ILI incidence to construct local forecasts with a 30-week horizon. The incidence data is constructed from a high-throughput syndromic surveillance system for feverish illness, consisting of a distributed network of thermometers with more than one million users across the US. These data are first used to quantify predictable variation in a city’s seasonal transmission pattern (“transmission templates”) based on historical data. We then use these transmission templates to produce long-range ILI forecasts across a range of scenarios for the upcoming influenza season, encompassing variation in city-specific reporting rates, susceptible depletion, and onset timing of the influenza season. We show that the best-in-ensemble forecast variants are skilled at a time horizon of 30 weeks, yet identifiable by strong performance in the early season. Our findings demonstrate the potential for distributed syndromic monitoring networks to generate reliable long-range ILI forecasts at the scale of individual cities, by leveraging high resolution ILI incidence data, combined with predictable variation among cities in seasonal transmission patterns.

## Materials and Methods

### Data

We use daily ILI incidence data for cities across the US, collected from a network of thermometers managed by Kinsa Inc. The network records temperature and approximate location of over one million de-identified users when they take their temperature (e.g. during an illness episode). The temperature readings are spatially aggregated and used to construct ILI time series that are highly correlated to Center for Disease Control and Prevention (CDC) ILI nationally (r > 0.95) and across CDC regions (r range 0.70-0.94) (*7*, *8*).

### Analysis

The analyses described here were conducted for nine US cities (see Figure 1). We estimated transmission templates using data from the 2016-2017 and 2018-2019 influenza seasons, and then forecasted ILI for the 2018-2019 season, starting Nov 1, 2018, with a horizon of 30 weeks. Our forecasting approach consists of the following steps.

**Figure 1.**
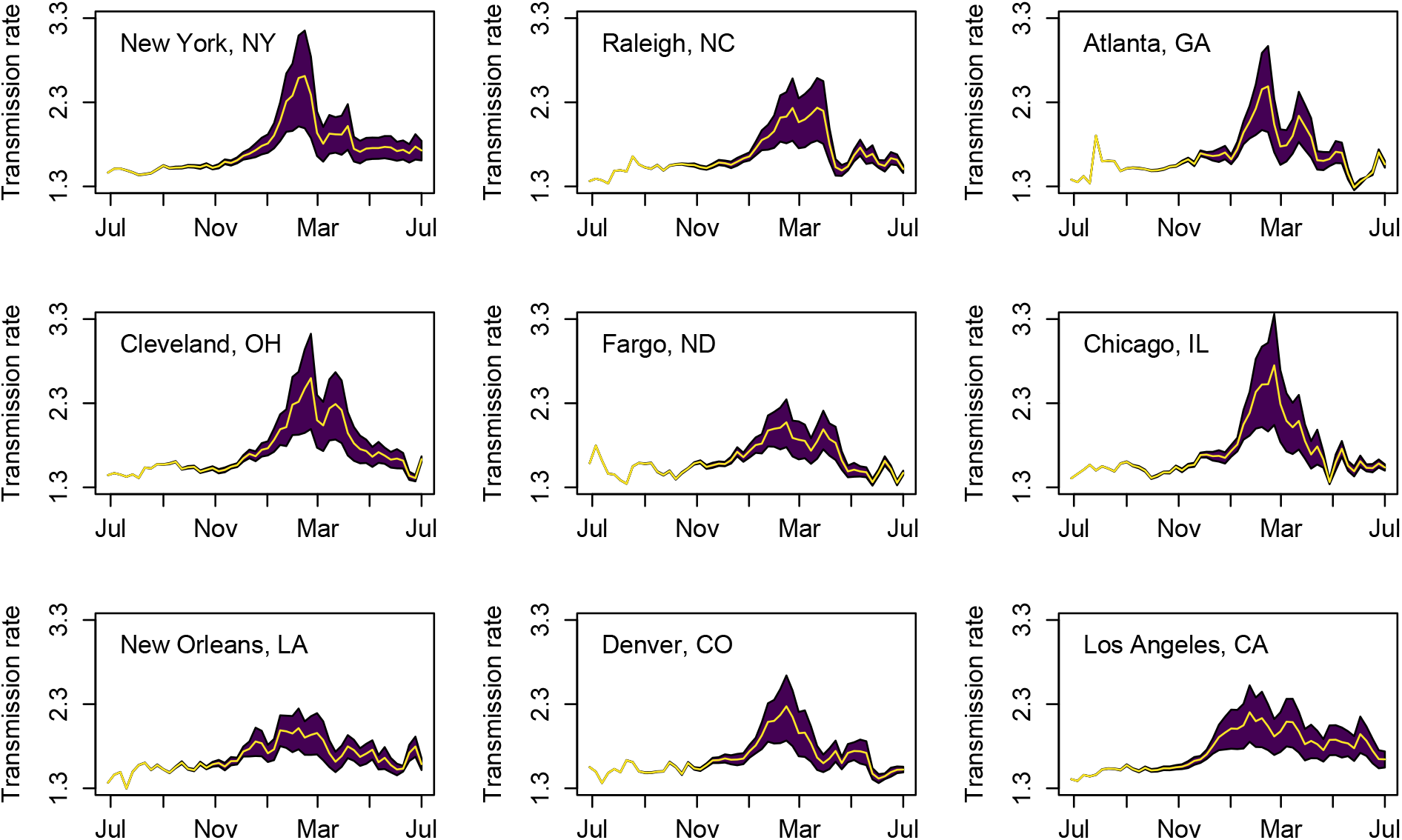
Seasonal patterns in transmission rates in nine cities (i.e. “transmission templates”), estimated using daily data from July 1, 2016 – Oct 31, 2018 (i.e., the 2016-2017, and 2017-2018 influenza seasons). Shaded regions enclose the mean weekly template values conditioned on α ranging from 1/3 to 3, which corresponds to peak susceptible depletion between of approximately 5-20%, with the yellow line indicated the mean value.

1. Starting with daily incidence data in a particular city *I_t_*,, where *t* represents time in days, use the data before the point of forecasting (Nov. 1, 2018) to estimate the daily reproductive number *R_t_* from (*9*) by solving

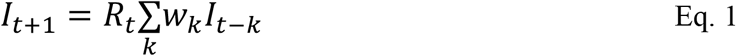

where *w_k_* is the infectivity profile for influenza, representing the probability that an individual who becomes infected on day *t* acquired the infection from an individual who became infected on day *t-k*. We approximate *w_k_* by an estimate of the generation time distribution for influenza, using a gamma distribution with a mean of 2.5 and variance of 0.7 days (*6*). This raw estimate of *R_t_* via equation (1) uses a smoothed version of *I_t_* using on a 30-day moving average. This smoothing removes high frequency noise, which, all else equal, prevents overfitting and thus improves forecasting performance. However, this smoothing can also alter the amplitude of seasonal fluctuations in *R_t_*, which can bias forecasts. To compensate we tune *R_t_* to 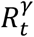, by selecting the scalar γ>0 to minimize squared error between *I_t_* and a “backcast” made by iterating equation 1 forward as

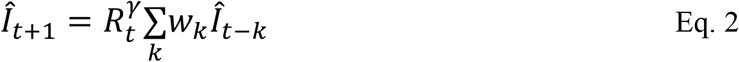

where 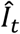 represents incidence forecasted t-steps ahead, and the objective function for minimization is

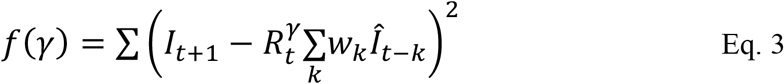

which typically yields values for γ between approximately 0.8 and 1.2, where γ=1 corresponds to no modification to the raw value for *R_t_*.
2. Estimate the fraction susceptible over time from cumulative incidence (*10*),

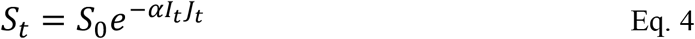

where *S*_0_ = 0.68 is the expected initial proportion susceptible (*11*), *J_t_* is equal to cumulative incidence since July 1 immediately prior to the current influenza season in the US (i.e. the season to be forecasted), and the reporting term *αI_t_* models the rate at which susceptibility declines with increasing cumulative incidence, adjusting for the rate false positives and under reporting. We thus make the simplifying assumption that reporting rates are linearly proportional to the current incidence *I_t_*, implying that as incidence increases a greater fraction of the true infected population is detected, and that a greater fraction of the cases that are detected are true positives. The resulting susceptible construction is conditioned on *α*, and different forecasting variants in the ensemble use different values of *α*.
3. Estimate the raw transmission rate as *β_t_ = R_t_/S_t_*, and the seasonal transmission template as a linear model of *β_t_* as a function of week of year. Let 〈*β*〉*_t_* represent the resulting transmission template – the expected transmission rate in a city in the week of year associated with day *t*. To accommodate annual differences in the onset of influenza season, we use forecast variants that apply a scalar shift parameter *τ*, such that 〈*β*〉*_t−τ_* is evaluated to calculate transmission rate at time *t*.
4. Forecast by propagating the model predictions forward using

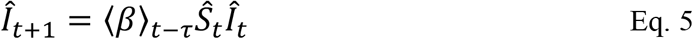 With forecasted susceptibility 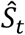 calculated using a modified version Eq. 4,

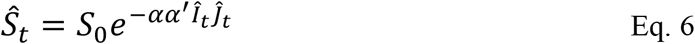

which includes a correction factor α’ for a systematic scalar deviation in susceptible depletion and reporting rate constants during the forecasted season (e.g. due to antigenic shift or variation in vaccination coverage).
5. Generate forecast variants by repeating steps 2-4 with different values of α and α’, as well as variations in a scalar template shift parameter τ. We then assess the skill of each variant in the ensemble over 30 weeks using root mean squared error (RMSE).

## Results

Seasonal transmission patterns appear to differ systematically among cities (Figure 1). This is consistent with previous work (*1*, *11*), which indicates these differences are driven by climate (specific humidity) as well as population size and density. The magnitude of these inter-city differences suggest they may be robust to underlying uncertainty in rates of susceptible depletion and reporting, represented by the polygons in Figure 1.

City-specific forecast ensembles always contained skillful variants at the 30-week time horizon (Figure 2). Ensembles were parameterized with these transmission templates but varied randomly in assumptions about reporting, susceptible depletion and the local onset of flu season. Both the best variant and the top 10% of variants performed well, as assessed by RMSE between observed and forecasted ILI incidence. The best variants are skilled enough to predict differences in the timing and number of peaks across cities (Figure 2).

**Figure 2.**
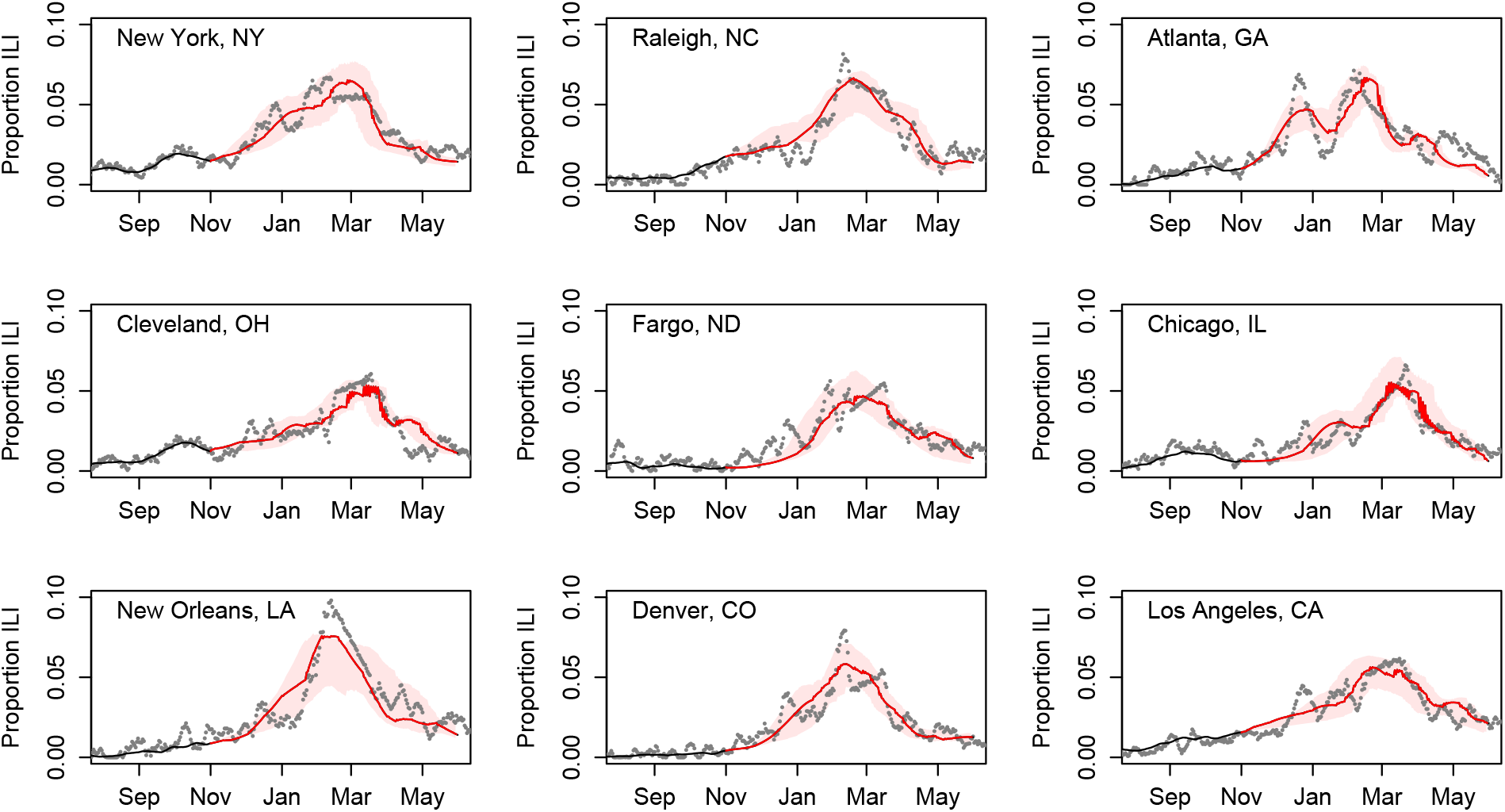
Forecasting the 2018-2019 influenza season 30 weeks ahead from Nov 1, 2018. Grey points show daily ILI incidence, the black line is abackcast, iterating equation 2 using estimated *R_t_* with the best fitting estimate of γ. The red line shows the “best-in-hindsight” variant, with skill assessed using RMSE on predicted versus observed incidence over the entire forecast. Red polygons enclose the best 10% of 1000 forecast variants which differ randomly in α, α’, and τ. Variants were created by sampling from uniform distribution with the following ranges: α ~ U(l/3, 3), α’ ~ U(l/2/,2), τ ~ U(−8 weeks, +8 weeks). Figure 3 shows the best variants are likely identifiable by their performance in the early season.

The best variants in hindsight consistently performed better than average throughout most of the season, starting within approximately 30 days of the forecast state date (Figure 3). This indicates the best variants could in principle be identified far in advance of peak incidence.

**Figure 3.**
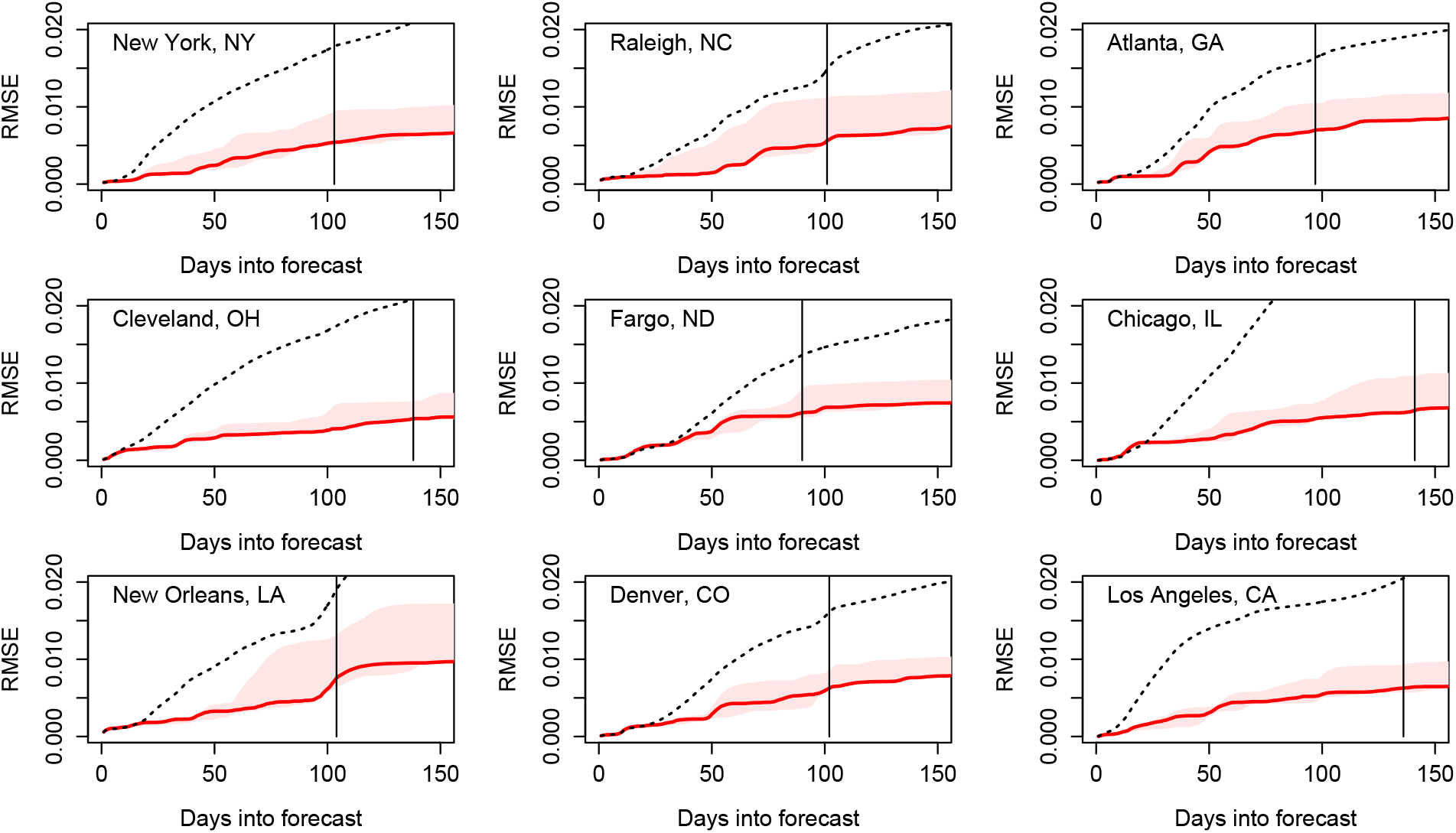
The best-in-hindsight variant (red line) has strong performance in the early season. Red polygons enclose the best 10% of 1000 forecast variants. Dashed line shows the mean performance across all forecast variants. Solid vertical line shows the timing of the peak incidence in the data for each city.

## Discussion

Long-range city-specific influenza forecasting requires integrating the impacts of local drivers of transmission, modeling local susceptible depletion and reporting, and anticipating the impacts of year-to-year variation in these processes. This interannual variation may be driven by secular demographic or climatic processes, random differences in regional spread patterns, antigenic evolution of influenza viruses, and ecological interactions between the respiratory pathogens that contribute to ILI incidence. The approach demonstrated here uses high-resolution incidence data to extract information on the average result of these processes across years, then generates an ensemble of forecasts to encompass year-to-year variation in ILI dynamics.

This path to long-range city-specific ILI forecasts rests on two results. First, we show that these forecast ensembles reliably contain a subset of highly skilled long-range forecasts (Figure 2). Second, we show that these forecasts can be identified early in the season (Figure 3). Early identification of skilled forecast variants should allow for enrichment of the ensemble, by replacing variants that are performing worse than average with variants similar to those that are performing better than average. Thus, the next step forward with this approach is developing and/or implementing methods for adapting and improving the forecasting ensemble as the influenza season progresses.

## Data Availability

Requests for data used in the manuscript can be made to Kinsa Inc.

## Author Contributions

IS conceived of and designed the Kinsa products, including its use as disease forecasting and monitoring system. SDC, PP, CAA, ALD and SDC designed and managed the real-time illness signal and geospatial platform. BDD developed the forecasting approach with feedback from SDC, PP, and IS. BDD wrote the first draft of the manuscript. All authors contributed to the final draft of the manuscript.

## Ethics Statement

An oversight determination for this research was issued by the Institutional Review Board of Oregon State University (Study Number IRB-2020-0687). The determination was that this research does not involve human subjects under the regulations set forth by Department of Health and Human Services 45 CFR 46. This work is not a clinical trial and is instead a population-level observational study, where all user information is anonymized and aggregated to the scale of United States cities. It therefore does not use individual, private or personally identifiable information.

## References

1. B. D. Dalziel et al., Urbanization and humidity shape the intensity of influenza epidemics in U.S. cities. Science. 362, 75–79 (2018).

2. C. M. Saad-Roy, A. B. McDermott, B. T. Grenfell, Dynamic perspectives on the search for a universal influenza vaccine. Journal of Infectious Diseases (2019), doi:10.1093/infdis/jiz044.

3. T. Bedford et al., Global circulation patterns of seasonal influenza viruses vary with antigenic drift. Nature. 523, 217–220 (2015).

4. J. D. Tamerius et al., Environmental Predictors of Seasonal Influenza Epidemics across Temperate and Tropical Climates. PLoS Pathog. 9 (2013), doi:10.1371/journal.ppat.1003194.

5. G. Brankston, L. Gitterman, Z. Hirji, C. Lemieux, M. Gardam, Transmission of influenza A in human beings. Lancet Infect Dis. 7, 257–265 (2007).

6. F. Carrat et al., Time lines of infection and disease in human influenza: a review of volunteer challenge studies. American Journal of Epidemiology. 167, 775–785 (2008).

7. A. C. Miller, I. Singh, E. Koehler, P. M. Polgreen, A Smartphone-Driven Thermometer Application for Real-time Population- and Individual-Level Influenza Surveillance. Clin. Infect. Dis. 67, 388–397 (2018).

8. A. C. Miller, R. A. Peterson, I. Singh, S. Pilewski, P. M. Polgreen, Improving State-Level Influenza Surveillance by Incorporating Real-Time Smartphone-Connected Thermometer Readings Across Different Geographic Domains. Open Forum Infectious Diseases. 6 (2019), doi:10.1093/ofid/ofz455.

9. A. Cori, N. M. Ferguson, C. Fraser, S. Cauchemez, A New Framework and Software to Estimate Time-Varying Reproduction Numbers During Epidemics. American Journal of Epidemiology. 178, 1505–1512 (2013).

10. D. E. te Beest, M. van Boven, M. Hooiveld, C. van den Dool, J. Wallinga, Driving Factors of Influenza Transmission in the Netherlands. American Journal of Epidemiology. 178, 1469–1477 (2013).

11. W. Yang, M. Lipsitch, J. Shaman, Inference of seasonal and pandemic influenza transmission dynamics. PNAS. 112, 2723–2728 (2015).

